# Determination of prevalence of HLA-B27 in patients with backache at Armed Forces Institute of Pathology (AFIP), Rawalpindi

**DOI:** 10.1101/2024.01.30.24302017

**Authors:** Abdul Rehman Haris, Furqan Maqsood, Hamid Nawaz Tipu, Dawood Ahmed

## Abstract

**Background:** HLA-B27 is a class I MHC protein that is associated with different diseases collectively called, spondyloarthropathies. It includes;

2. Ankylosing spondylitis, which causes inflammation of the bones in your spine
3. Reactive arthritis, which causes inflammation of your joints, urethra, and eyes, and in some cases lesions on your skin
4. Juvenile rheumatoid arthritis
5. Anterior Uveitis, which causes swelling and irritation in the middle layer of your eye

**Objectives:** To determine the prevalence of HLA-B27 in patients with backache at the Armed Forces Institute of Pathology in Rawalpindi. This descriptive cross-sectional study was conducted between June 2020 and Nov 2020.

**Material and methods:** Of the 243 patients, 167 males and 76 females were tested for HLA-B27 during the study period. Venous blood samples (3 mL) were collected in EDTA tubes and processed for HLA-B27 gene identification on the cell surface. A two-color flow cytometry panel was used to analyze samples using a BD FACS Canto II flow cytometer on BD FACS Diva software. The dot plot was created using an isotype control in four quadrants, and positive and negative populations of cells were identified.

**Results:** A total of 243 patients were analyzed for the presence of HLA-B27 using flow cytometry. Of the 243 patients, 167 were male and 76 were female patient.51 male patients tested positive, with a positivity rate of 30.53% among the male population and 20.98% among the general population. In contrast to the male population, 12 female patients tested positive for the HLA-B27 gene, with a positivity rate of 15.78% among the female population and 4.93% among the general population. The total positivity rate was 25.92%, with a higher prevalence in the male population included in this study.

**Conclusion:** All individuals tested for HLA B27 were included in this study. This study showed that positivity rate of HLA-B27 is more in males with a percentage of 20.98% as compared to female population (4.93%).

## Introduction

Human leukocyte antigens (HLA) are proteins that present antigens to T cells. MHC proteins are alloantigens (i.e., they differ among members of the same species). In humans, these proteins are encoded by Major Histocompatibility complex (MHC) genes on the short arm of chromosome 6. Three of these genes (HLA-A, HLA-B, and HLA-C) encode class I MHC proteins. Several HLA-D loci encode class II MHC proteins (HLA-DP, HLA-DQ, and HLA-DR). MHC class I proteins are found on the surface of all nucleated cells, with some exceptions (i.e., astrocytes present in the CNS), whereas MHC class II proteins are found on the surface of professional antigen-presenting cells (APCs) such as macrophages, dendritic cells, and B-cells. The complete class I protein is composed of a 45,000 molecular weight heavy chain that is non-covalently bound to β_2_ microglobulin. The main function of class I molecules is to present endogenous antigen peptides to CD8+ cytotoxic T-cells’ class II which are composed of two heavy chains, alpha and beta chains, which are non-covalently bound. Their function is to present processed antigenic peptides derived from exogenous sources to CD4+ helper T cells. In addition to class I and II MHC, there are class III MHC proteins”. Unlike other MHC, such as class I and class II, whose structure and function are well defined, MHC class III is poorly defined structurally and functionally. Gene clusters were present between Classes I and II. MHC class III includes several proteins with immune functions, such as complement (C2 and C4), cytokines (TNFs), and heat shock proteins. These proteins are synthesized in the liver and extrahepatic mononuclear phagocytes (1).

### HLA-B27

Although MHC proteins protect the body from harm and help the immune system to identify the differences between healthy body tissues and foreign substances, some MHC proteins contribute to immune system dysfunction. HLA B-27 is one of the proteins associated with various diseases. HLA-B27 is strongly associated with ankylosing spondylitis (AS) and other associated inflammatory diseases referred to as “spondyloarthropathies,” including Psoriasis, Ankylosing spondylitis, inflammatory bowel disease, and reactive arthritis.

### Structure and subtypes

Human leukocyte antigen (HLA) B27 is a class I surface antigen encoded by the B locus in the significant histocompatibility complex (MHC) on chromosome 6 and presents antigenic peptides (obtained from self- and non-self-antigens) to T cells. HLA-B27 has a high degree of hereditary polymorphism, with 105 known subtypes, named HLA-B*27:01–HLA-B*27:106, encoded by 132 alleles. The most well-known subtypes related with ankylosing spondylitis are HLA-B*27:05 (Caucasians), HLA-B*27:04 (Chinese), and HLA-B*27:02 (Mediterranean population). In Chinese populations, HLA-B*27:04 is related to a more noteworthy ankylosing spondylitis hazard than HLA-B*27:05. Two subtypes, HLA-B27*06 and HLA-B27*09, appear to have no association with the illness (2).

In the local population, there are a wide number of HLA-B*27 subtypes and an increased frequency of the B*2707 allele in patients with AS. The B* 2706 allele appears to have a protective role in the population studied because it was found in healthy controls. HLA-B*27:03 and 07 are the predominant subtypes in Punjabis and Pathans, respectively (3).

### Disease association

The presence of HLA-B27 is related to certain immune system disorders, including

- Ankylosing spondylitis, which causes inflammation of the bones in your spine
- Reactive arthritis, which causes inflammation of your joints, urethra, and eyes, and in some cases lesions on your skin
- Juvenile rheumatoid arthritis
- Anterior Uveitis, which causes swelling and irritation in the middle layer of your eye Doctors may advise the HLA-B27 test to monitor the progression of these autoimmune diseases.

### Worldwide distribution

Commonness of HLA-B27 shifts, especially for everyone. For instance, approximately 8% of Caucasians, 4% of North Africans, 2-9% of Chinese, and 0.1-0.5% of people of Japanese plunge have the gene that codes for this antigen (4).

### Study objectives

This study aimed to determine the prevalence of HLA-B27 in patients with backache at the Armed Forces Institute of Pathology (AFIP), Rawalpindi.

The objective of our study was to determine the prevalence of HLA-B27 in patients with backache at AFIP in Rawalpindi.

### Historical Perspective

HLA-B27 is one of the most interesting molecules in the field of medicine. Its contribution to the etiopathogenesis of spondyloarthropathies and other diseases and its protective mechanism in certain infections continues to challenge our understanding of its immunological and physiological roles. Evidence has shown that it reduces the virulence of HIV and other infections (5). Since its first prominence in 1973 with the discovery of its intimate association with ankylosing spondylitis (6), much has been learned about its immunobiology and involvement in both etiopathogenesis and disease prophylaxis.

The natural function of HLA-B27 is to form a complex with β_2_ macroglobulin, which then binds to short antigenic peptides derived from intracellular antigens and presents them to cytotoxic T cells, which then kill the infected cells (7). The ‘arthritogenic peptide’ hypothesis proposes that particular properties of the HLA-B27 peptide binding groove to bind and present the antigen peptides can exhibit molecular mimicry with specific arthritogenic self-peptides. This process can also initiate cross-reactivity with self-peptides. This autoimmune reaction may lead to chronic inflammation (8). HLA-B27 also appears to complement Salmonella invasion in intestinal epithelial cells (9). It is impossible to find live bacteria or DNA in joints; however, fragments of Chlamydia, Yersinia, and Salmonella have occasionally been found in patients (10). Although HLA-B27 is associated with different diseases, the exact mechanism of disease development is still not clear and requires further investigation.

Human leukocyte antigen (HLA) B27 is a class I surface antigen encoded by the B locus in the significant histocompatibility complex (MHC) on chromosome 6 and presents antigenic peptides (obtained from self- and non-self-antigens) to T cells. HLA-B27 is strongly associated with ankylosing spondylitis (AS) and other associated inflammatory diseases referred to as “spondyloarthropathies,” including Psoriasis, Ankylosing spondylitis, inflammatory bowel disease, and reactive arthritis.

Komsalova et al, 2020 conducted a prospective study titled “Predictive values of inflammatory back pain, positive HLA-B27 antigen and acute and chronic magnetic resonance changes in early diagnosis of spondylarthritis. Study of 133 patients” at Hospital Marina Salud, Denia, Alicante, Espana. In the study population, 41.4% of patients had inflammatory back pain more often in patients with spondylarthritis and 83% vs. 18.6% SpA/not SpA patients. Of these, 30.3% were positive for HLA-B27 and 49% vs. 20% for SpA/non-SpA patients (11).

Hamid Nawaz Tipu et al, 2017 conducted cross sectional study titled “HLA B27 prevalence among patients of Spondyloarthropathies’ Armed Forces Institute of Pathology, Rawalpindi. Of the 329 studies, 252 were males (76.6%) and 77 females (23.4%). Of these 329 cases, 77 (23.4%) tested positive for HLA B27. The positive samples included 66 males (26.2%) and 11 females (14.3%) (12).

Nelly Raymond Ziade et al., 2017 performed a literature review on 27 studies performed between 1978 to 2012 titled “HLA B27 in Middle Eastern and Arab countries: systemic review of strength of association with axial spondylarthritis and methodological gaps”. They reported that the prevalence of HLA B27 in the general population of Oman is (0.3%) and Turkey (6.8%). The prevalence of HLA B27 in axial spondylarthritis is 26.2% in Lebanon and 91% in Turkey. The prevalence of HLA B27 in all spondylarthritis cases ranged from 13.87% (Lebanon) to 69.4% (Kuwaiti). Peripheral spondylarthritis is less associated with HLA B27 than axial spondylarthritis, indicating that there is a need to differentiate between these two to calculate the prevalence of HLA B27. Eight of the 27 studies revealed that the prevalence of HLA B27 ranged from 21.63% (Morocco) to 105.6% (Syria). All these differences between the results may be due to different study methods, sample sizes, classification criteria, and absence of control groups (13).

John D et al., 2013 conducted a study titled “The epidemiology of back pain, axial spondylarthritis and HLA B27 in United States” at Division of Rheumatology, The University of Texas Health Science Center at Houston (JDR), Houston, Texas and Division of Rheumatology, Cedras-Cenai Medical Center (MHW), Los Angeles, California. According to the NHANES 2009, the age-adjusted prevalence of HLA B27 in the U.S was 6.1%. HLA B27 was prevalent in 7.5% of non-Hispanic whites and 3.5% of all other U.S. races and ethnicities. In Mexican Americans, the prevalence of HLA B27 was 4.6%. In the Blacks group, the presence of HLA B27 was not significant (1.1%). For age 50-69 the prevalence of HLA B27 was 3.6%, suggesting that the frequency of HLA B27 decreases with increasing age. Compared to the older population, the prevalence of HLA B27 in the younger population of the U.S (age 20-49) was 7.3%. This study showed that there is a significantly lower prevalence of HLA B27 in the older population than in the younger population (14).

Adrian B. Beckingsale., 1984 conducted a study titled “Acute anterior uveitis, ankylosing spondylitis, back pain and HLA B27 at University of Leicester School of M medicine, Leicester. The study population consisted of 169 patients. Of these 169 patients, 76 (44.97%) were known HLA B27 positive cases and 93 (55.02%) were HLA B27 negative cases. Back pain and symptoms of ankylosing spondylitis were found in 46 (60.52%) of these 76 positive patients (44.73% (34) males and 15.78% (12) females), and 13 patients whose HLA B27 was negative (61.53% (8) males and 38.46% (5) females) also showed symptoms of back pain and ankylosing spondylitis (15)

Ivo Jajic et al., 1979 conducted a study titled “The Role of HLA B27 in Diagnosis of Low Back Pain” at ward for Rheumatic Disease, Department of Orthopedics, Medical Faculty, Zagreb University, Yugoslavia. He investigated 652 patients with back pain to determine the HLA B27 status in these individuals. Of these, 42.33% were positive for HLA B27 antigen. Of these positive patients, 128 had ankylosing spondylitis, and the remaining 148 did not show any signs or symptoms of ankylosing spondylitis. Of the 276 patients positive for HLA B27, 204 were male and 72 were female. Of the 128 patients with ankylosing spondylitis 114 were men and 14 were women. Another 376 subjects tested negative for HLA B27, but showed other non-specific HLA antigens (16).

## Material and methods

This study used a descriptive cross-sectional design. A Non-Probability Universal Sampling Technique was used to collect samples. The duration was six months, from June 2020 to Nov 2020. This study was performed at the Immunology Department, Armed Force Institute of Pathology, Rawalpindi. Training Institute of Armed Forces Post Graduate Medical Institute (AFPGMI) Rawalpindi. All patients with a history of back pain were recruited at the Armed Forces Institute of Pathology (AFIP) Rawalpindi for the evaluation of HLA-B27. A total of 243 samples were analysed during the study period.

### Ethical Consideration

Institutional consent was obtained from the institutional ethics committee of the Armed Forces Institute of Pathology (AFIP), Rawalpindi. The results were included to keep them confidential and strictly for academic purposes. Clinical specimens were assigned specific identification numbers to maintain their anonymity.

### Statical analysis

Data were analyzed using the Statistical Package for Social Sciences (SPSS) version 21.0. Data were presented as frequencies, percentages, mean, and standard deviations. The confidence level of the study was kept at 95%, hence a ‘‘p’’ value less than 0.05 indicated a significant association

### Procedure protocol

A total of 243 patients were tested for HLA-B27 in the Immunology Department during the study period. Three ml venous blood was collected in EDTA tubes and tested for HLA-B27. A two-color flow cytometer panel was used to analyze the samples. First, the cells were stained with fluorescein isothiocyanate (FITC) and phycoerythrin (PE)-labeled monoclonal antibodies and incubated for 30 min in the dark at room temperature. RBCs were lysed by adding the BD FACS lysing solution. The remaining cells were washed with phosphate-buffered saline (PBS) and fixed in formaldehyde (3.3% formaldehyde prepared in PBS). Samples were analyzed with a BD FACS Canto II flow cytometer using the BD FACS Diva software with a count of 20,000. The cell population was identified using forward and side scatter and gated for further analysis. The dot plot was created using an isotype control in four quadrants, and positive and negative populations of cells identified.

**Figure 1:**
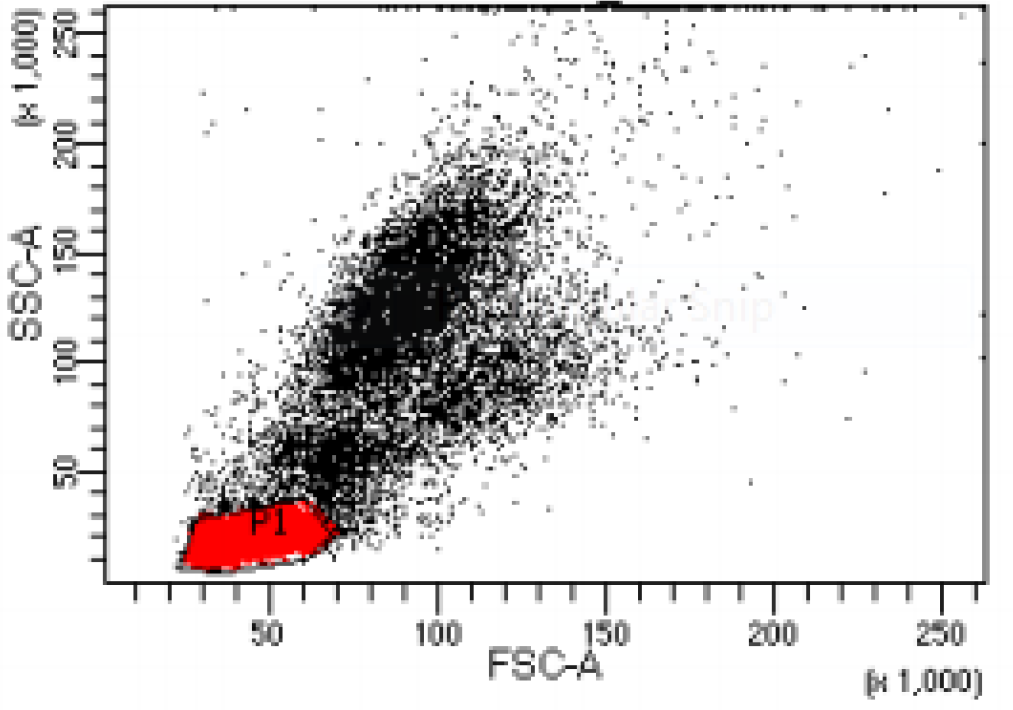
Dot plot shwere owing gated population of cells.

**Figure 2:**
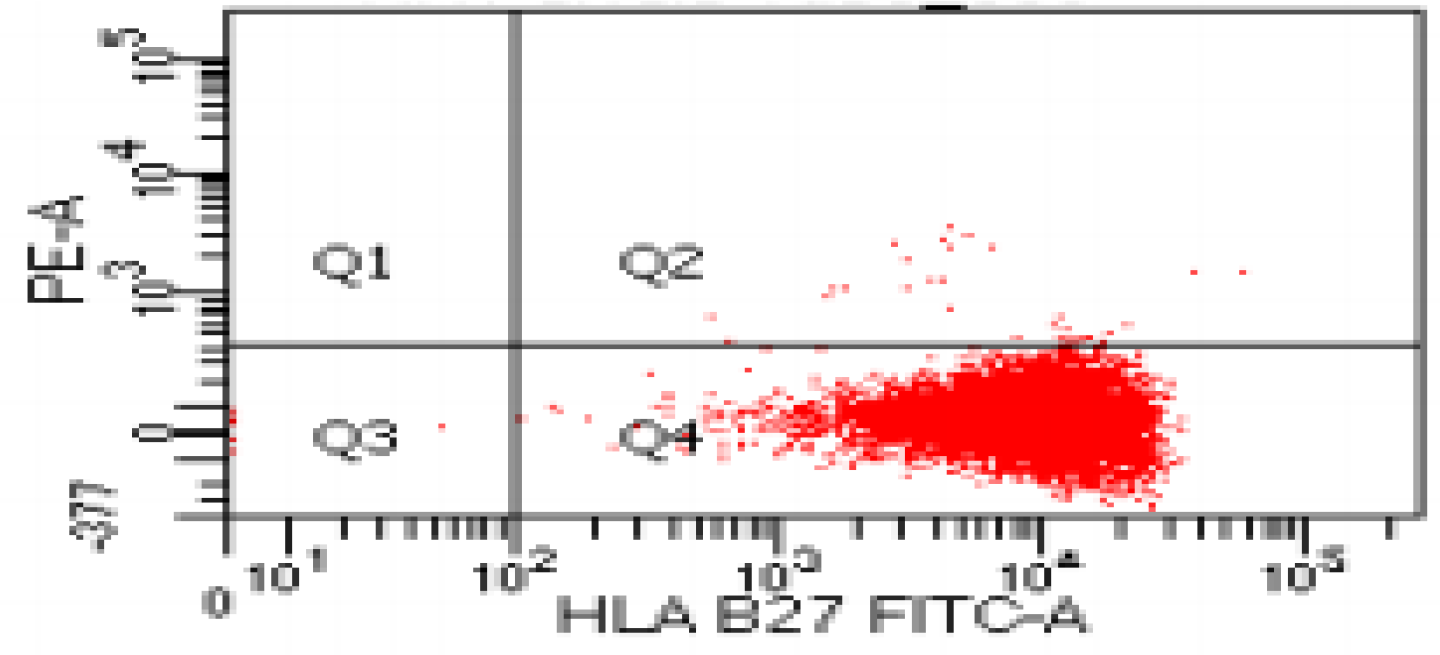
Dot plot showing positive population of cells for HLA B27.

**Figure 3:**
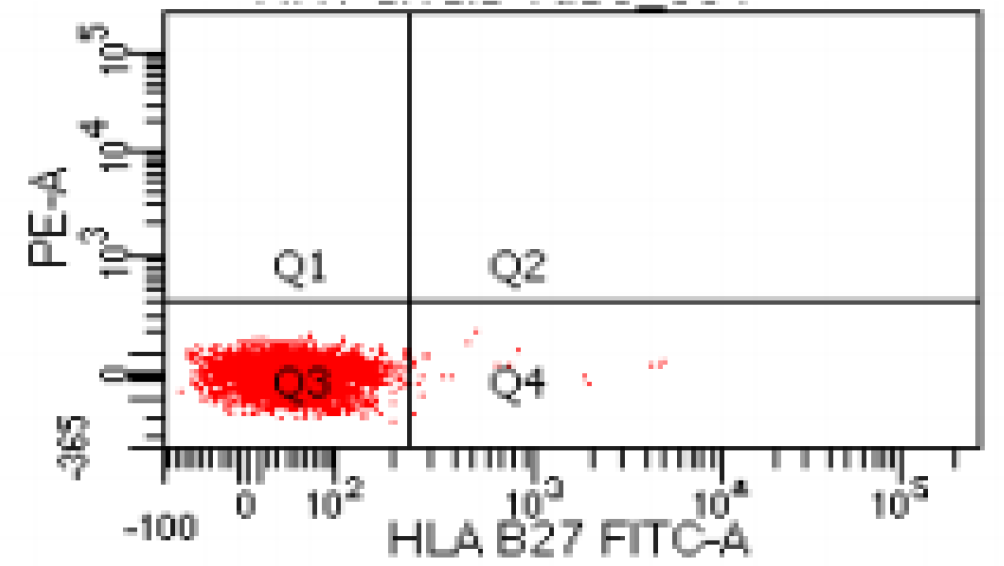
Dot plot showing negative population of cells for HLA B27.

## Results

A total of 243 patients were analyzed for the presence of HLA-B27 using flow cytometry. Of the 243 patients, 167 (68.72%) were male and 76 (31.27%) were female, with ages ranging from 8 to 82 years (mean age 34.79 ± 12). 51 male patients tested positive, with a positivity rate of 30.53% in the male population and 20.98% in the general population. In contrast to the male population, 12 female patients tested positive for the HLA-B27 gene, with a positivity rate of 15.78% among the female population and 4.93% among the general population. The total positivity rate was 25.92%, with a higher prevalence among males.

**Table 1:** Age wise frequency distribution of HLA B27.

| Age | Frequency | Positive | Negative |
| --- | --- | --- | --- |
| 8-15 | 13 | 02 | 11 |
| 16-23 | 22 | 06 | 16 |
| 24-31 | 59 | 17 | 42 |
| 32-39 | 79 | 24 | 55 |
| 40-47 | 34 | 09 | 25 |
| 48-55 | 24 | 04 | 20 |
| 56-63 | 07 | 01 | 06 |
| 64-71 | 02 | 00 | 02 |
| 72-79 | 02 | 00 | 02 |
| 80-87 | 01 | 00 | 01 |
| | $\Sigma f=243$ | $\Sigma x_1=63$ | $\Sigma x_2=180$ |

**Figure 4:**
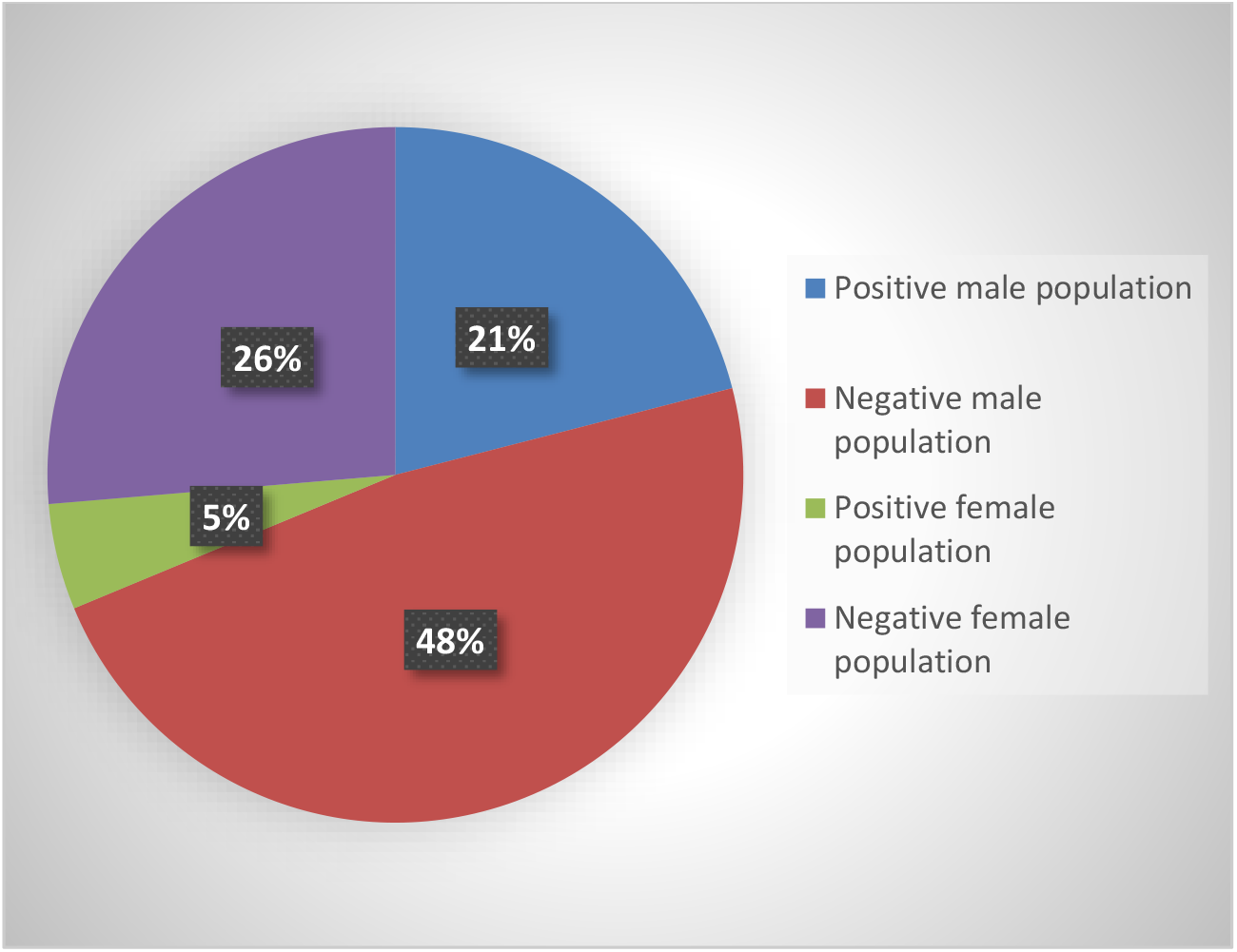
Gender wise distribution of HLA B27.

**Figure 5:**
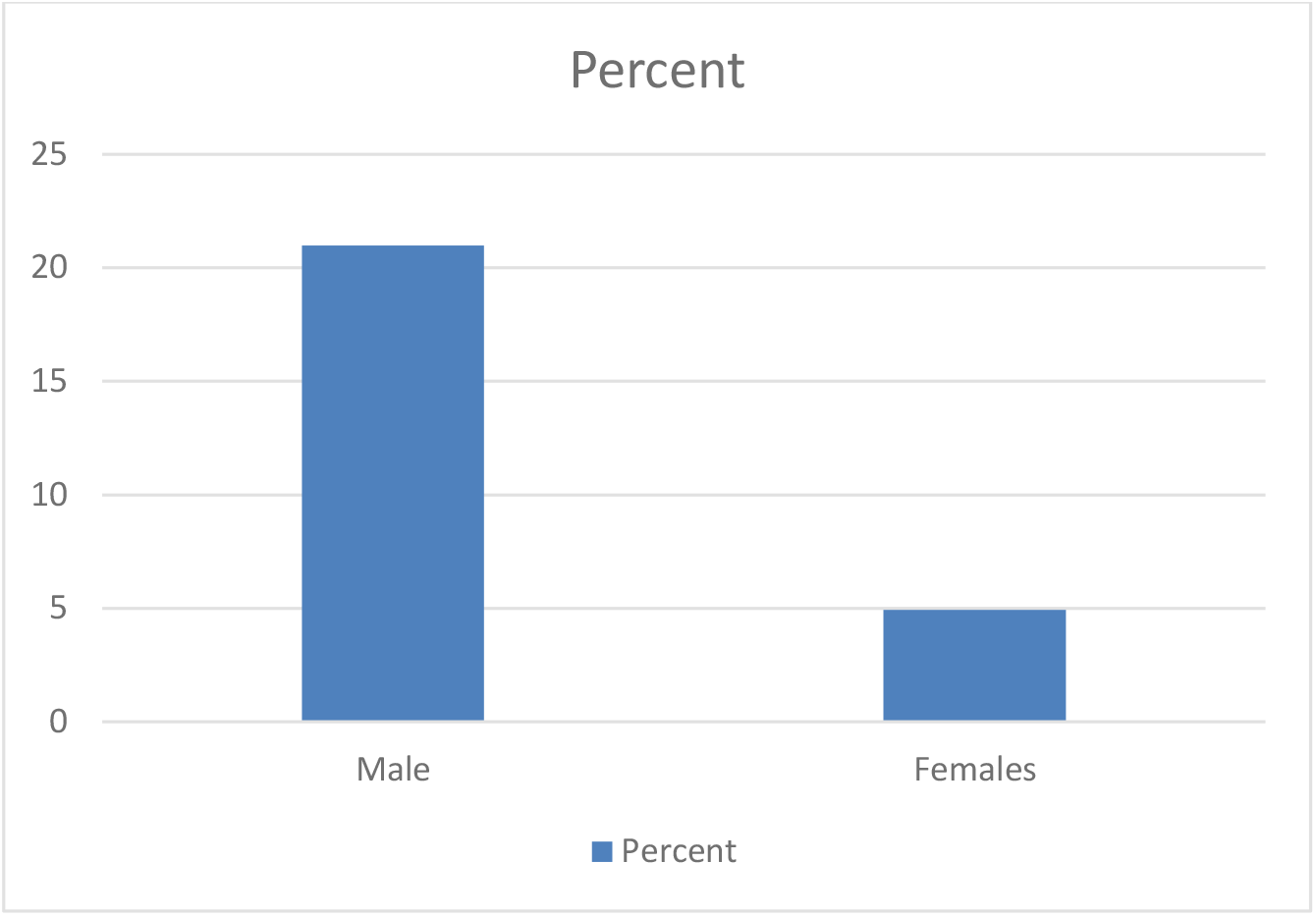
Males vs. females’ prevalence of HLA B27.

## Discussion

Human Leukocyte Antigen (HLA) B27 is a class I MHC molecule consisting of an alpha chain noncovalently bound to β_2_ microglobulin. It is highly polymorphic, with 105 known subtypes, named HLA-B*27:01–HLA-B*27:106. Individuals with the HLA B27 gene are at a higher risk of developing autoimmune diseases because they are associated with some diseases. These diseases are collectively called “Spondyloarthropathies (SpA).” These disorders include psoriatic arthritis, reactive arthritis, inflammatory bowel disease, juvenile idiopathic arthritis, and ankylosing spondylitis (AS). Ankylosing spondylitis is thought to be associated with HLA B27.

Worldwide prevalence of HLA B27 is about 8% of Caucasians, 4% of North Africans, 2-9% of Chinese, and 0.1-0.5% of people of Japanese plunge have the gene that codes for this antigen (2).

J.T Gran et al., 1984 conducted a study titled “Prevalence of HLA-B27 in Northern Norway. *Scandinavian journal of rheumatology”*. HLA-B27 incidence has been studied in 176 consecutive blood donors in Tromsø, Northern Norway. The prevalence of HLA-B27 was 15.9%, which was significantly higher than that reported in Southern Norway (10%). The frequency of 15.9% is comparable to the prevalence of this antigen found in Northern Sweden (16.6%) and Finland (14–16%) (17).

Abdelrahman et al. (2012) conducted a study titled” Prevalence of HLA-B27 with ankylosing spondylitis in Qatar” in the Rheumatology section, Department of Medicine, Hammad General Hospital, Doha, Qatar. His study population included 66 Arabs, 52 Asians (Indians, Pakistan, Bengalis, and Iran), and one Western (Irish). *Of these*, 82 were positive (69%) for HLA-B27. Among Arabs, 49/66 were positive (74%) were positive. Among Asians, 32/52 were positive (61%) were positive. Furthermore, 9 Qatari patients (10 males and one female) were positive (82%), 14/19 Jordanians/Palestinians were positive, and 9/10 (90%) Egyptians were positive. Among Asians, 19/26 Indians were positive (73%), similar to Arabs (18).

Hamid Nawaz Tipu et al, 2017 conducted cross sectional study titled “HLA B27 prevalence among patients of Spondyloarthropathies’ at Armed Forces Institute of Pathology, Rawalpindi. They reported that the prevalence of HLA B27 was 23.4%, with a higher positivity rate in the male population than in the female population (12).

In the present study, the positivity rate of HLA B27 was 25.92%, with a higher prevalence in the male population (20.98 %). The age group of 24 to 44 years is at a higher risk of developing these diseases, with a higher prevalence of the HLA B27 gene.

## Conclusion

HLA B27 is a class I molecule that is associated with various diseases. It is a highly polymorphic gene, with 105 subtypes. These diseases include psoriatic arthritis, juvenile rheumatoid arthritis, inflammatory bowel disease, and ankylosing spondylitis (AS). HLA B27 was significantly positive in patients with a history of backache. Techniques, such as flow cytometry and PCR, can be used to evaluate HLA B27. Radiographs of patients with back problems can also be examined for evaluation. The positivity rate of HLA B27 varies in different regions of the world depending on genetic factors, sex, and age.

## Study limitation

The limitation that could influence the result can be; The population that was included is limited to AFIP only so, results cannot be applied to the whole Pakistani population

## Data Availability

All data produced in the present study are available upon reasonable request to the authors

